# Environmentally persistent free radicals in household dust: the seasonal and longitudinal trends

**DOI:** 10.1101/2023.10.22.23297366

**Authors:** Dwan Vilcins, Prakash Dangal, Slawomir Lomnicki, Stephania Cormier, Wen Ray Lee, Peter D Sly

## Abstract

**Objective:** Epidemiological links between air pollution and adverse health outcomes are strong, but the mechanism(s) remain obscure. A newly recognised combustion by-product, environmentally persistent free radicals (EPFRs), may be the missing link. EPFRs persist for extended periods of time in the environment, however very little is known about the presence of EPFRs inside homes where prolonged exposure is likely to occur. The objective of this study is to explore the presence of EPFRs in household dust and ascertain if EPFR concentration is stable across time and season.

**Material and methods:** The ORChID/ELLF cohort is a longitudinal birth cohort (n=82) with dust samples collected from the family vacuum cleaner at multiple time points. EPFR characteristics were assessed with electron paramagnetic resonance. Our team developed an algorithm to estimate oxygen-weighted concentration and impact score for risk of adverse health outcomes. Kruskal-Wallis rank sum test and Fisher’s exact tests were used to assess seasonal differences. A simple mixed-effects linear regression, with random intercepts on participant ID, was employed for longitudinal analysis of EPFR concentration in households that did not move.

**Results:** 83 participants returned 238 dust samples. EPFRs were measured in virtually all samples. EPFR concentration was stable across visits, when controlling for season and ambient air pollution (p=0.05), Oxygen-weighted EPFRs were also stable. There was a seasonal trend, with concentration (p=<0.01), oxygen weighted concentration (p=<0.01) and g factor (p=0.05) all significantly lower in summer months.

**Conclusion:** Our results indicate that the concentration of EPFRs in household dust are stable across time in households that did not move, but the oxygen-centred radicals are more sensitive to changes. These findings suggest that exposure to EPFRs occurs in the home and may be a significant place for exposure to highly biologically reactive EPFRs.

## 1. Introduction

Air pollution is globally recognized as a threat to human health (1). Advances in scientific investigation have enabled high quality research showing air pollution is harmful to human health even at low exposure levels (2, 3). In response, the World Health Organization released their latest air quality guidelines in 2021, which lowers the recommended 24-hour and annual limits for ambient pollutants such as particulate matter, nitrogen dioxide and ozone to a significantly lower threshold (4). In terms of human exposure to pollutants, ambient air pollution is only part of the story. Indoor generation of air pollutants adds to overall exposure and sometimes occur at higher rates than ambient air pollutants (5). Exposure to high levels of indoor air pollutants are well known in environments that rely heavily on solid fuel as an energy source (6). Populations living in countries with relatively good ambient air quality can find their indoor exposure to pollutants is higher than from ambient air, and is compounded by sealed environments that do not allow regular air exchange (7). Indoor air pollution can be generated from residential activities such as cooking (8), burning incense or candles (9), cigarette smoking (10), biomass burning for cooking or heating (11) or inadequate ventilation (7), and further raised by the infiltration of ambient air pollutants (12).

A major pollutant of concern is particulate matter (PM), most notably fine particles less than 2.5 microns in diameter (PM_2.5_). A mature body of evidence supports the association of PM exposure with a range of adverse health effects, including higher mortality (3), asthma (2), decrements in lung function (13) and adverse birth outcomes (14). PM_2.5_ is a complex structure, that typically contains a carbon core with pollutants such as metals, hydrocarbons, sulphur, and phenols adsorbed onto the surface (15). The exact components of each particle differ by source. The components of PM may drive the health effects, with previous work finding coronary heart disease is associated with the sulphur component of PM and cerebrovascular disease is associated with organic carbon (15). This work highlights the importance of understanding the components of PM, in addition to total PM mass, to which populations are exposed. A component of increasing concern is environmentally persistent free radicals (EPFRs). EPFRs were first discovered in the 1950s, but it was in the 2000’s when it was discovered that exposure to EPFRs can occur through environmental carriers such as air and soil (16). EPFRs are formed within the cool zone of combustion processes, and in the presence of a transition metal oxide (17). It is the relationship between the transition metal and organic species on the particles that allows the formation of EPFRs (17). The lifetime of an EPFR is dependent on the PM components involved, and ranges from half an hour to several years (18). As well as the presence of EPFRs, electron spin resonance allows the exploration of the speciation of EPFRs; that is, whether the EPFR shows signals of carbon or oxygen centre, and any additional elemental signatures (16). It has previously been shown that EPFR speciation may be dependent on environmental conditions such as season (19).

A proposed mechanism underlying the negative health impacts of air pollution exposure is oxidative stress. Exposure to air pollution can trigger the generation of reactive oxygen species, which in turn induces oxidative stress and damages DNA (20). The formation and source of ROS are still debatable and recent evidence suggests that ROS can be generated from EPFRs found on PM_2.5_ (20-22). EPFRs, therefore, may be a key part of the causal pathway between exposure and outcome. The evidence for their presence in human environments remains in its infancy, and mostly focuses on their presence in air (17). As PM deposits into the indoor environment these EPFRs can settle into household dust and remain present for extended periods of time. Therefore, household dust can be a key driver of long-term exposure to EPFRs. Children living in households with a high EPFR concentration in dust are more likely to report wheezing (23). What is unknown is whether the concentration and speciation of EPFRs is stable across time in households, or whether they depend heavily on ambient environmental conditions. The aim of this study is to explore to what extent EPFRs are present in household dust; whether EPFRs in dust have a relationship with season; and whether the concentration and speciation of EPFRs present in household dust is stable across time.

## 2. Methods

### 2.1. Participants

The Early Life Lung Function (ELLF) study is an extension of the Observational Research in Childhood Infectious Diseases study (ORChID); a prospective, community-based longitudinal study of children recruited during pregnancy and followed until two years of age. Children who completed ORChID were invited to participate in the ELLF extension project. The ELLF study ran from April 2014 to December 2019 and aimed to explore the impact of household and environmental exposures in early childhood on respiratory health. Children attended an annual study visit, with 82 children enrolled into ELLF and 79 completing the 7 year follow-up. The study was extended after the 7 year visits to include home visits. 50 families were invited to participate in two home visits: one during winter and one during summer. 24 families agreed to participate and completed at least one home visit during the study period between January 2020 and July 2022. The Children’s Health Queensland Human Research Ethics Committee reviewed and gave ethical approval for the study (HREC/13/QRCH/156).

### 2.2. Household dust collection

Household dust samples were scheduled for collection at the 3, 4, 6, and 7 year follow-up visits, and the summer and winter home visits. Participants in the 3-7 year follow-up were requested to bring a sample of their vacuum cleaner dust to their study visit at the clinic, which was then frozen at -22°C. Participants in the home visits were asked to vacuum their home and not empty the bag at least 24 hours prior to the home visit. The research team collected the dust from the household vacuum during the home visit and transported it to the centre on ice, where it was frozen on arrival. Dust samples were shipped to Louisiana State University, and EPFRs were analysed using electron paramagnetic resonance.

### 2.3. EPFR outcome measures

The EPFR g factor is the ratio of an electron’s magnetic moment to its angular momentum and indicates the nature of the paramagnetic species (17). The g factor indicates whether the free radicals contain more oxygen centre characteristics, which indicates a higher level of biological activity. The g factor was used as a continuous variable.

The concentration of EPFRs within house dust was measured as the number of spins / gram. We further calculated the concentration of oxygen-weighted EPFRs (O-EPFRs) within the dust sample. The correction for the content of oxygen centered radicals in the samples was based on the analysis of the EPR spectra parameters, using a linear combination of the 3^rd^ power of the g-tensor shift in respect to the position of the pure oxygen centered radical (2.0049) and spectral broadening represented by the value of delta H peak-to-peak. The resulting adjustment factor was then multiplied by the spin-concentration number. O-EPFRs are more likely to be biological active and participate in the redox system.

### 2.4. Covariates

Year of collection and season of collection were extracted from the date the participants reported collecting their dust sample, or the date the research team collected the sample for participants in home visits. Ambient air pollution was calculated for each household at each time point of the study using a satellite-based, land-use regression models (Sat-LUR) (24) that we developed and validated and which is described in previous studies (25) (26). The Sat-LURs were developed for annual mean concentration of particulate matter ≤ 2.5 μm (*PM*_*2*.*5*_), using satellite, land use and other spatial predictors. The indicative spatial resolution of the models is up to 100 m in urban areas and up to 500 m in rural and remote areas (27). This model captured between 52% and 63% of variability in PM^2.5^, with an RMSE of 1 to 1.2 μg/m^3^ (28). Participant’s residential addresses were geocoded to 6 decimal places.

### 2.5. Statistical analysis

Dust samples with an insufficient weight could not be analysed for EPFRs and were removed from analysis. Samples with an EPFR concentration below the limit of detection were given 1/2 the value of the lowest concentration (1.61e+13 spins/g). This method was chosen as the data was skewed, even under a log transformation (29). Data was cleaned and summarised using standard descriptive techniques. Households were allocated to move vs. did not move during the study period on a strict criterion requiring no move at any time point. Correlation between variables was tested using Pearson Product-Moment Correlation. The unpaired two samples Wilcoxon test with continuity correction was used to assess difference between seasonal EPFR concentration and g factor. Households that returned at least two samples and did not move were included in the longitudinal analysis. A mixed-effects linear regression was performed, with random intercepts on participant ID to examine if visit time point (year) was significantly associated with g factor, concentration, and O-EPFRs. A second mixed-effects model was employed to control for effects of season and ambient PM_2.5._ Estimated marginal means were calculated for pairwise contrasts by study year, and correction for multiple comparisons was performed using the Tukey method.

## 3. Results

### 3.1. Descriptive summary

246 individual dust samples were provided from 83 participants during the study period. Eight samples had insufficient weight for analysis and 11 samples were below the limit of detection and thus received imputed concentration values. In total, this paper reports on 238 dust samples from 83 participants. The 3 year visit and 7 year visit had the highest number of participants returning a dust sample, with very low numbers for the 4 year (*n=*24) and 6 year (*n*= 30) visits. There were 22 samples in summer and 24 samples in winter from participants in the home visit arm of the study. No participants provided dust samples for all time points. When limiting to those participants who had at least 2 timepoints available and did not move, there were 55 records available for longitudinal analysis. Figure 1 shows an overview of the samples.

**Figure 1:**
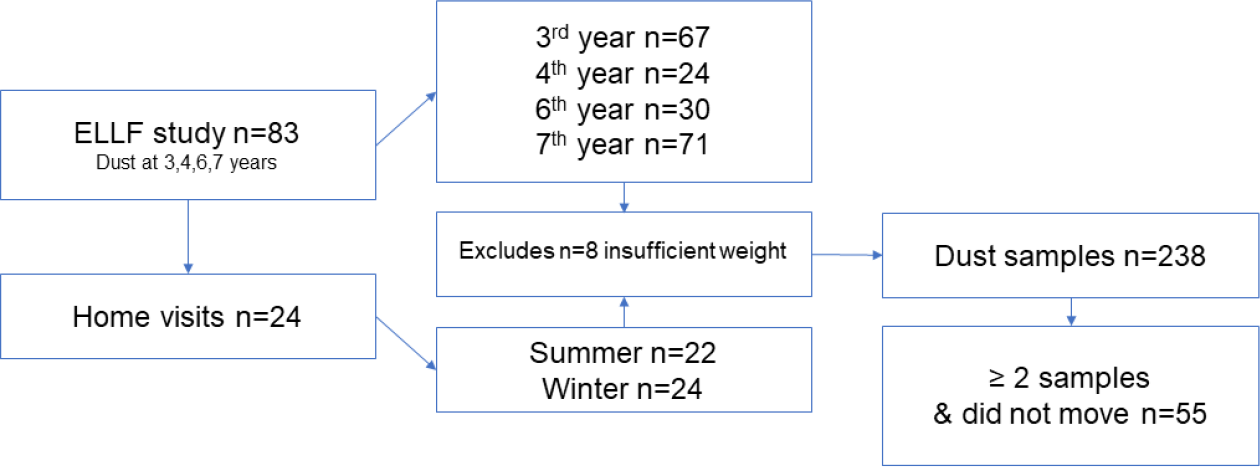
Overview of sample flow from the ELLF study and follow-up home visit study.

EPFRs were found in all participant households (*n*=83, 100%), and 95.5% of the dust samples contained measurable EPFRs. A summary of the EPFR outcomes is provided in Table 1. The median g factor across all samples was 2.003 gauss (Q1,Q3 [2.003e+00, 2.004e+00]). The median concentration was 3.120e+17 spins/g (Q1,Q3 [1.772e+17, 4.898e+17]), with a median O-EPFR of 1.500e+17 spins/g (Q1,Q3 [7.725e+16, 2.718e+17]).

**Table 1:** Description of household dust samples (n=238), collected from n=83 participants in Brisbane, Australia.

| Variable |  | Summary |
| --- | --- | --- |
| g factor (median [IQR]) |  | 2.003e+00 [2.003e+00, 2.004e+00] |
| Number of dust samples |  | 238 |
| Concentration of EPFRs spins/g (median [IQR]) |  | 3.120e+17 [1.772e+17, 4.898e+17] |
| Oxygen-weighted concentration of EPFRs (median [IQR]) |  | 1.500e+17 [7.725e+16, 2.718e+17] |
| Visit of collection (%) | 3<sup>rd</sup> Year | 67 (28.2) |
|  | 4<sup>th</sup> Year | 24 (10.1) |
|  | 6<sup>th</sup> year | 30 (12.6) |
|  | 7<sup>th</sup> year | 71 (29.8) |
|  | Summer | 22 (9.2) |
|  | Winter | 24 (10.1) |
| Moved during study period (%) | Yes | 176 (73.9) |
|  | No | 62 (26.1) |

A correlation was present between g factor and year of dust collection. Season was weakly correlated with concentration and oxygen weighted concentration. Ambient annual PM_2.5_ was not correlated with any of the EPFR measures in household dust. The correlation matrix is presented in the supplementary materials – Figure S1.

### 3.2. Seasonal trends in EPFRs

#### 3.2.1. G Factor

The g factor of EPFRs in household dust had a seasonal pattern. The lowest g factor was seen in summer months (median g factor summer=2.0031 gauss (IQR 0.0005), rising across spring (2.0033 gauss (IQR 0.0006)), winter (2.0033 gauss (IQR 0.0006)), and highest in autumn (2.0034 gauss (IQR 0.0005)). Pairwise comparisons showed statistically significant differences between summer with the colder seasons (winter (*P=*0.02) and autumn (*P=*0.02) but not with spring (*P=*0.27). When restricting only to households that did not move, the differences between warm and cold seasons became more pronounced. Pairwise comparisons (with Bonferroni correction) found significant differences between spring and winter (*P*=0.01) and spring and autumn (*P*=0.03), in addition to significant differences between summer with both autumn (*P*=<0.01) and winter (*P*=<0.01).

#### 3.2.2. Concentration

There was a seasonal element to EPFR concentration in house dust, with concentration being lowest in the summer months (median concentration in summer=2.28e+17 spins/g (IQR 2.3e+17), winter=3.4e+17 spins/g (IQR 3.2e+17), autumn=3.63e+17 spins/g (IQR 2.2e+17), spring=4.24e+17 spins/g (IQR 3.6e+17) (Figure 2). EPFR concentration was significantly lower in summer compared to all other seasons (Kruskal-Wallis rank sum test, *H* (3) = 16.4, *P* = <0.001). There was no difference between the other seasons in pairwise comparisons. When restricting analysis to homes that did not move in the study period there was a small change in median EPFR concentration in each season, but the overall trend remained the same.

**Figure 2:**
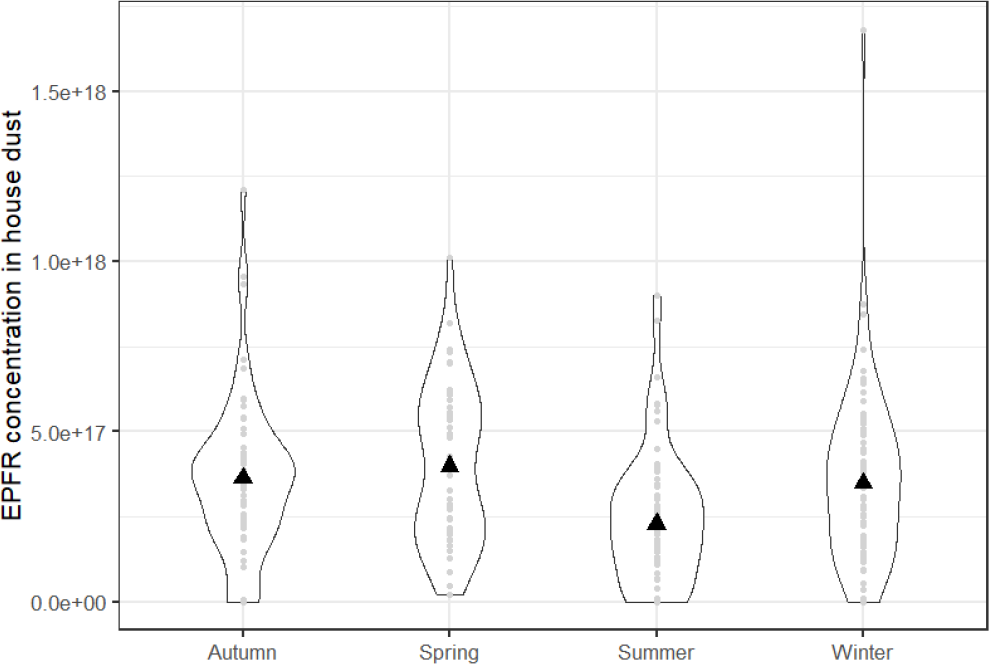
EPFR concentration in house dust by season (two homes with outliers greater than 4E+18 spins/g were removed from this figure).

#### 3.2.3. Oxygen-weighted concentration (O-EPFR)

The O-EPFR in house dust followed the same pattern as concentration (Figure 3), with the lowest value seen in summer. The median O-EPFR in summer was 9.2e+16 spins/g (IQR 1.2e+17), winter= 1.6e+17spins/g (IQR 2.1e+17), spring= 1.8e+17 (IQR 1.9e+17) with autumn showing the highest values (1.8e+17 spins/g (IQR 2.1e+17)). There was a significant difference between O-EPFR in summer compared with all other seasons (Pairwise comparisons using Wilcoxon rank sum test with continuity correction summer vs. winter *P*=0.007; vs. autumn *P*=0.007; vs. spring *P*=0.01). Restricting analysis to those homes who did not move during the study period did not change the results.

**Figure 3:**
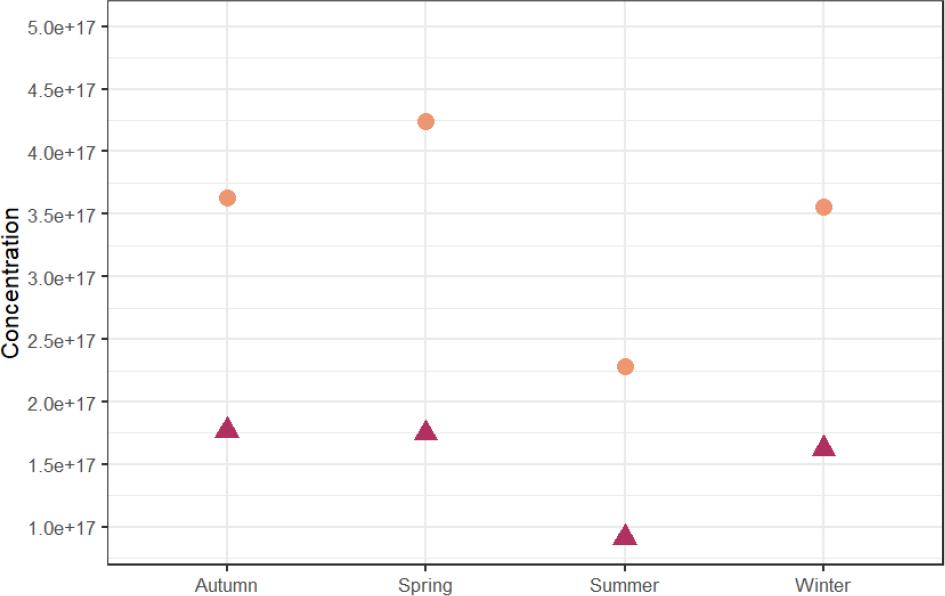
Median EPFR concentration (orange circles) and median oxygen-weighted EPFR concentration (red triangles) from household dust in Brisbane, Australia.

### 3.3. Longitudinal trends

The primary outcome of this study is to ascertain whether the concentration and/or speciation of EPFRs in household dust is stable over time. To explore the stability of EPFRs we limited analysis to households that returned a minimum of two dust samples and did not move during the study period. EPFR characteristics by year are shown in supplementary table S1.

#### 3.3.1. Stability of EPFR g factor in household dust

The median g factor was higher in the first two years of dust collection and dropped after 2015. The g factor of EPFRs showed statistically significant differences across the study years. These differences were predominately for 2014 and 2015 compared to the later study years. There were significant differences in the g factor across years 2017-2022. Analysis of longitudinal trends using a mixed-effects model, and controlling for household level factors, season of dust collection and ambient PM_2.5_ supported the finding of significant differences in g factor across the years, but driven by the higher g factor in 2014 and 2015.

#### 3.3.2. Stability in the concentration of EPFRs in household dust

Stability of EPFR concentration was first examined in an unadjusted analysis which found some significant differences between years. Controlling for household level confounding and season fully attenuated these differences in collection year. The inclusion of ambient PM_2.5_ did not change the results. This suggests that EPFR concentration is stable across time when accounting for household level differences and season (Table 2).

**Table 2:**
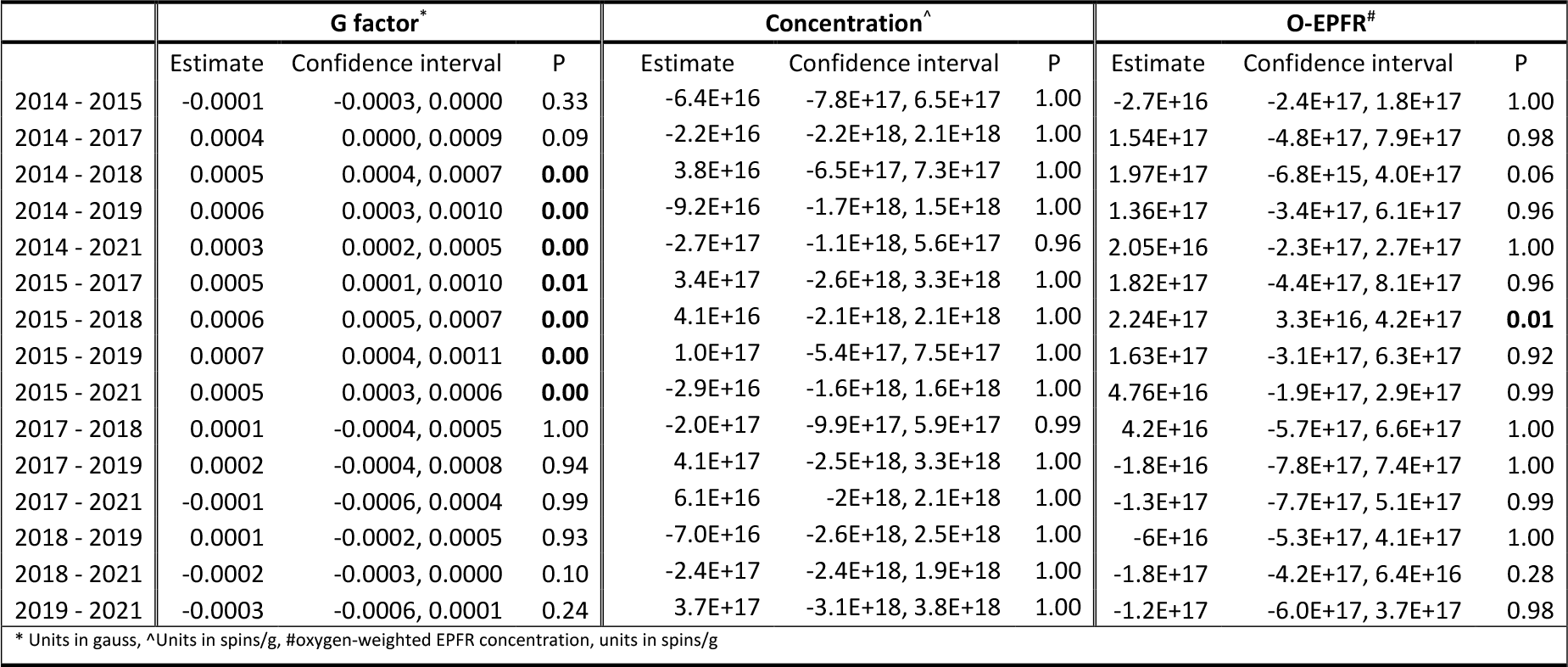
Estimated means for EPFR concentration and speciation assessing the stability over longitudinal collection of household dust from a Brisbane, Australia cohort of children. Models adjusted for household level factors, season and ambient PM_2.5_.

#### 3.3.3. Oxygen-weighted concentration

Unadjusted, pairwise analysis of the O-EPFRs found variations across several years, which largely attenuated in the mixed-effects models controlling for household level differences and season. Controlling for ambient PM_2.5_ almost completely accounted for changes across the study year. The one exception is a significant difference between 2019 and 2015, with a borderline difference between 2014 and 2018. All other pairwise comparisons show no significant differences, suggesting that presence and amount of O-EPFRs are stable in household dust after accounting for differences in households, season and ambient PM_2.5_ (Table 2).

## 4. Discussion

In this longitudinal cohort, we found measurable EPFRs in household dust for every participant. This finding shows that exposure to EPFRs in the home is ubiquitous, and not limited to airborne exposures. Given that airborne EPFRs depend on the lifetime of particles (17), exposure in household dust may represent a more persistent environmental exposure medium. We found a strong seasonal relationship with EPFRs, which was largely driven by summer months exhibiting lower EPFR concentrations. A seasonal relationship with EPFRs has previously been shown (31, 32). The concentration of EPFRs associated with airborne PM_2.5_ in Beijing, China were lower in the summer months, which was attributed to clearing of PM_2.5_ by precipitation and the influence of ambient temperature (33). In a separate region of China (Wanzhou), the same seasonal pattern of lower concentration of airborne EPFRs in hotter months was seen, although the higher EPFR concentration in autumn and winter was attributed to straw burning and coal burning, respectively (34). Further, a correlation analysis found that secondary processes involved in EPFR formation may be more important in winter (34). We found a seasonal pattern for g factor in our study, which was also seen in previous studies (31, 34), however it should be noted that unlike seasonal patterns on concentration which is similar across studies. The variation in g factor by seasonal is not stable across the different studies and may be driven by local factors. As far as we know, we are the first team outside of China to explore the seasonal patterns of EPFRs and the first to explore seasonal trends of EPFRs in household dust. Our work lends further credence to the role of season influencing EPFR concentration and speciation. In our main analysis we found that the concentration of EPFRs and O-EPFRs is stable across the study years. This is an important finding, as it allows us to gain an understanding of the potential long-term exposure, where repeat sampling is not possible. To our knowledge we are the first team to assess the presence of EPFRs in household dust and the longitudinal exposure to EPFRs with repeated measures in the same environment across many years.

The presence of EPFRs in air (31-34), fly ash (16), soils and sediments (35) and road dust (36) has been previously reported, however household dust is an important medium due to its persistence in households. Children tend to be exposed to toxicants in dust more than adults due to behaviours unique to children such as more time on the floor and hand-to-mouth behaviours (37). Further, households closer to dust generating sources such as busy roads and incinerators have more dust infiltration (38).

The evidence that EPFRs have a role in adverse health outcomes is growing. Several pre-clinical studies found EPFRs on PM_2.5_ can trigger cardiac and pulmonary diseases through generation of reactive oxygen species in rats (39-42). An EPFR-like compound was also found to induce pulmonary and cardiovascular issues in animals. In adult rodent models of asthma were found to have increased oxidative stress and immune responses in the lungs (43). EPFRs demonstrated to trigger inflammation and hyper-responsiveness in the airway (40) and worsen influenza infection with results showing higher morbidity and decreased survival rate in neonatal rats (44). A study of EPFRs and wheeze in the ELLF cohort has previously found that children living in homes with categorised as having a high exposure to EPFRs had higher persistent wheezing (58.3%) and more current wheezing symptoms (52.7%) compared to children living in the low EPFR exposure group (23). This body of work demonstrated the importance of studying the role of EPFRs as a causal mechanism between air pollution and adverse health effects.

The longitudinal nature of this study and the repeated measurements of EPFRs in household dust over many years is a strength of this study. Our ability to match modelled air quality data for each residential address created a unique dataset for the analysis of EPFRs. Our team is highly experienced in the measurement of EPFRs. However, some limitations remain. The small sample size did not allow us to perform more sophisticated models to determine the source of the EPFRs. Our modelled air quality is provided as an annual average which gave us a measure of long-term air quality and not seasonal variation. Human exposure to EPFRs is emerging as a key contaminant of concern and future research should be conducted to further explore the role of indoor generators of EPFRs as a source of exposure.

## 5. Conclusion

Indoor exposure to EPFRs present in household dust is ubiquitous, even in environments with relatively good air quality. There is a seasonal variation in the concentration of EPFRs in house dust, but the concentration across time is stable.

## Data Availability

Data are available from the cohort after review and approval by the study investigators

## Notes

### Competing Interest Statement

The authors have declared no competing interest.

### Funding Statement

This work was supported by the National Institute of
Environmental Health Sciences, USA (3P42ES013648-08A1S1)

### Author Declarations

The Childrens Health Queensland Human Research Ethics Committee reviewed and gave ethical approval for the study (HREC/13/QRCH/156).

