## Supplementary materials for "Environmentally persistent free radicals in household dust: the seasonal and longitudinal trends"

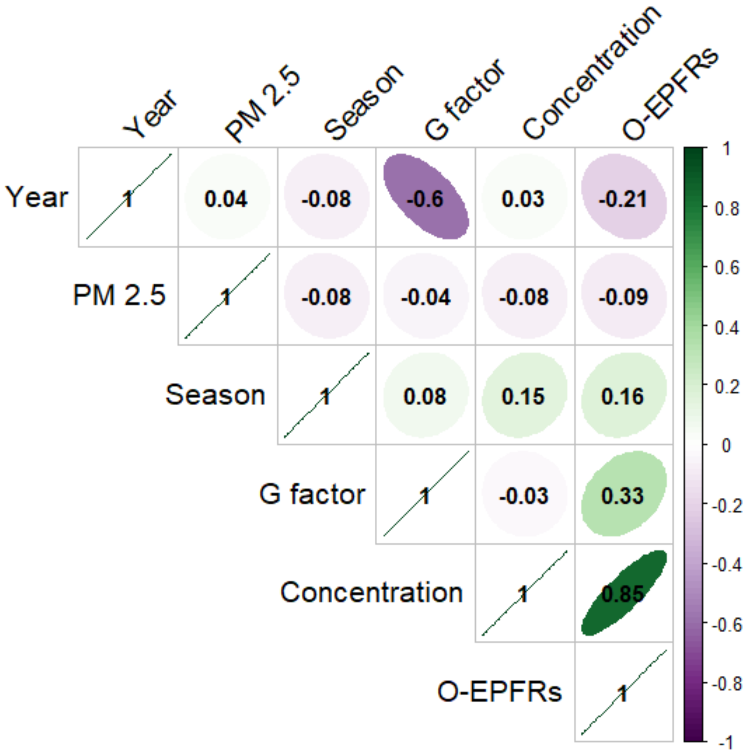


Figure S1: Matrix showing Pearson Product-Moment Correlation coefficient between EPFR outcomes from household dust and predictors. Year and season are based on date of dust collection, $PM_{2.5}$ is annual ambient values, O-EPFRs are the oxygen-weighted concentration of EPFRs in household dust.

Table S1: EPFR characteristics by year of collection for household dust samples collected from the ELLF study participants who returned at least two samples and did not move during the study period.

| | 2014 | 2015 | 2017 | 2018 | 2019 | 2021 | 2022 |
|---|---|---|---|---|---|---|---|
| **Number of dust samples** | 29 | 35 | 2 | 51 | 21 | 21 | 13 |
| **g factor (median [IQR])** | 2.0036 [2.0035, 2.0037] | 2.0038 [2.0036, 2.0038] | 2.0030 [2.0029, 2.0030] | 2.0031 [2.0030, 2.0032] | 2.0031 [2.0029, 2.0032] | 2.0032 [2.0031, 2.0033] | 2.0032 [2.0031, 2.0036] |
| **EPFR concentration (median [IQR])** | 5.340e+17 [4.2e+17, 6.15e+17] | 4.140e+17 [3.405e+17, 5.395e+17] | 2.810e+17 [2.795e+17, 2.825e+17] | 2.180e+17 [1.27e+17, 3.08e+17] | 2.880e+17 [1.97e+17, 3.72e+17] | 3.560e+17 [1.19e+17, 5.1e+17] | 1.610e+13 [8.05e+12, 1.16e+17] |
| **0-weighted concentration (median [IQR])** | 3.050e+17 [2.43e+17, 3.92e+17] | 3.080e+17 [2.585e+17, 3.755e+17] | 9.965e+16 [9.248e+16, 1.068e+17] | 8.320e+16 [4.91e+16, 1.36e+17] | 1.120e+17 [8.06e+16, 1.43e+17] | 1.365e+17 [7.875e+16, 2.082e+17] | 5.440e+16 [4.125e+14, 8.54e+16] |
